# Gender Differences in Socioeconomic Inequalities in Body Mass Index and Health-Related Mediators Among Young and Early Midlife Finns

**DOI:** 10.64898/2026.11.10.26367699

**Authors:** Emma Järvensivu, Lari Järvensivu, Morris Viitanen, Ossi Rahkonen, Tea Lallukka

**Affiliations:** Department of Public Health, University of Helsinki, Helsinki, Finland

**Author notes:** Corresponding author: Emma Järvensivu. Lari Järvensivu, Morris Viitanen, Ossi Rahkonen, Tea Lallukka.

## Abstract

**Introduction:** Socioeconomic differences in body mass index (BMI) are well established, but less is known about gender differences and their behavioral pathways. We examined gender differences in the association between socioeconomic position (SEP) and BMI, and whether dietary factors and physical activity mediated the associations.

**Methods:** We used survey data from the employees of the City of Helsinki aged 19–39 collected in 2017 (n = 4,400, 80% women). SEP was measured by highest educational attainment, and the examined mediators were consumption of fruit and vegetables, red and processed meat, sweets and confectionery and alcohol as well as leisure-time vigorous physical activity (VPA) and sedentary time. We conducted gender-specific multiple mediation analyses using structural equation modeling and additional analyses using multiple imputation by chained equations.

**Results:** With increasing educational attainment, mean BMI decreased from 26.4 kg/m² to 24.0 kg/m² among women, and from 26.6 kg/m² to 25.2 kg/m² among men. Lower SEP was associated with lower consumption of fruit and vegetables, higher consumption of red and processed meat, and lower VPA in both genders. Women reported healthier diets but less VPA than men. The strongest SEP–BMI mediator among women was VPA (indirect effect: 0.35, 95% confidence interval (CI) 0.26, 0.44), followed by fruit and vegetable consumption (0.15, 95% CI 0.08, 0.23) and red and processed meat consumption (0.06, 95% CI 0.02, 0.11). Among men, significant mediators were red and processed meat consumption (0.12, 95% CI 0.03, 0.23), sedentary time (0.11, 95% CI 0.01, 0.24) and VPA (0.10, 95% CI 0.02, 0.21).

**Conclusions:** Socioeconomic inequalities in BMI appear stronger among women and are shaped through partly different mechanisms between genders. Improving VPA may be relevant among low- SEP women, whereas reducing sedentary time may be more relevant among low-SEP men. Reducing red and processed meat is beneficial for both genders.

## Introduction

Overweight and obesity are rising globally and in Finland, and in high-income countries, higher body weights are increasingly concentrated in lower socioeconomic groups [1]. According to the World Health Organization, in 2022 the global prevalence of overweight and obesity among adults aged 18 years and older was 43% and 16%, respectively [2]. In Finland, the prevalence of obesity has increased substantially between 2000 and 2023, rising from 21% to 30% among men and from 22% to 30% among women, with the most pronounced increases seen among adults aged 30–44 years [3]. However, recent trends suggest that in high income countries, the rise in obesity is plateauing for instance due to better affordability of healthier foods compared to low- and middle-income countries [4].

Socioeconomic position (SEP) refers to social and economic differences within society that arise from educational, occupational, economic, and cultural resources [5]. Education is a widely used indicator of SEP because it is established relatively early in adulthood and captures resources that may shape health knowledge, working conditions, material circumstances, and health-related behaviors [6]. In high-income countries, body mass index (BMI) generally shows an inverse socioeconomic gradient, although the strength of this association differs by gender [7]. Previous studies have often reported stronger and more consistent inverse associations between SEP and obesity among women than among men [8,9]. This pattern was already identified in the Finnish population in the 20 century [10] and has since been replicated in later national and international studies [8,11].

Dietary behaviors are among the most plausible mediators linking SEP to later BMI. Lower SEP is linked to consuming fewer fruits and vegetables [12,13]. Fruit and vegetable consumption can support weight regulation through lower energy density, higher fiber content and greater satiety, and previous research suggests a higher consumption is associated with reduced obesity [14]. By contrast, red and processed meats are often energy dense and embedded in dietary patterns associated with greater adiposity but have also been independently associated with weight gain [15]. The consumption is higher in those with lower SEP, which makes it a potential mediator of the SEP– BMI association [12]. Sweets and confectionery are also plausible mediators, although the strongest evidence linking sugar intake to obesity concerns sugar-sweetened beverages [16]. Alcohol should also be considered, since it can contribute to excess energy intake directly or indirectly by weakening dietary restraint. The epidemiological evidence on alcohol and adiposity is mixed, but heavier intake and some beverage patterns seem less favorable [17]. However, no clear socioeconomic differences have been observed in sweets and confectionery or alcohol consumption, although the gender pattern suggests that women consume more sweets, whereas men consume more alcohol [12].

In addition to diet including alcohol consumption, moderate-to-vigorous physical activity as well as sedentary time are potential mediators since they are socioeconomically patterned, potentially modifiable and consistently linked to weight management [18–20]. Physical activity and less sedentary time may influence body weight not only through energy expenditure but also through improved appetite regulation and satiety signaling [21]. However, physical activity is socially patterned, and these patterns vary by domain and intensity: higher socioeconomic groups tend to be more active during leisure time and to accumulate more high-intensity physical activity, whereas lower socioeconomic groups tend to accumulate more occupational and light-intensity physical activity [19,20,22]. Sedentary time is positively associated with both general and abdominal obesity, and although sedentary time contributes to energy balance, the evidence is unclear whether increased sedentary time leads to increased BMI or whether the associations are due to reverse causation [23–25]. Socioeconomic differences also vary by sedentary domain, with higher SEP groups tending to report more occupational sitting, while lower SEP groups tend to report more leisure-time sitting [26].

Despite extensive research on socioeconomic differences in obesity, SEP–gender interactions in the associations are rarely explored. More specifically, the mechanisms through which SEP contributes to the development of obesity, and whether these mechanisms differ by gender, remain insufficiently understood. We hypothesize that lower SEP is linked to a higher BMI partly through dietary factors and physical activity and that the absolute effects of the mediators are larger among women because the SEP–obesity gradient is steeper in women. First, we examined gender differences in the associations between SEP and BMI. Second, we examined whether dietary factors and physical activity similarly mediate associations among both genders.

## Methods

### Data and study population

This study is part of the Helsinki Health Study, which examines the health and well-being of employees of the City of Helsinki, the largest employer in Finland with approximately 38,000 employees across diverse occupational sectors [27,28]. The target population consisted of 11,459 employees born in 1978 or later (aged 19–39 years in 2017) who had been employed for at least four months with a contract of at least 50% regular working hours. Data collection occurred in autumn 2017 (Phase 1) via online and mailed questionnaires, supplemented by shorter telephone interviews for non-responders.

The initial response rate was 51.5% (n = 5,898). While non-response analysis indicated slightly higher participation among those with higher SEP, the sample was found to reasonably represent the target population [27]. We excluded telephone interviewees (n = 510) because the interviews did not include all variables of interest needed for this study. We also excluded those with missing data on education, BMI as well as one or more of the mediators, as shown in Figure 1. Gender had no missing values. Outliers in physical activity and sedentary time were identified as values more than three standard deviations from the mean, following previous studies [20]. The final analytical sample included 4,400 participants of whom 80% were women, reflecting the gender distribution of both the City of Helsinki and the broader Finnish municipal sector. Two-thirds of the participants were aged 30–39 years, and one-third was aged 19–29 years.

**Figure 1.**
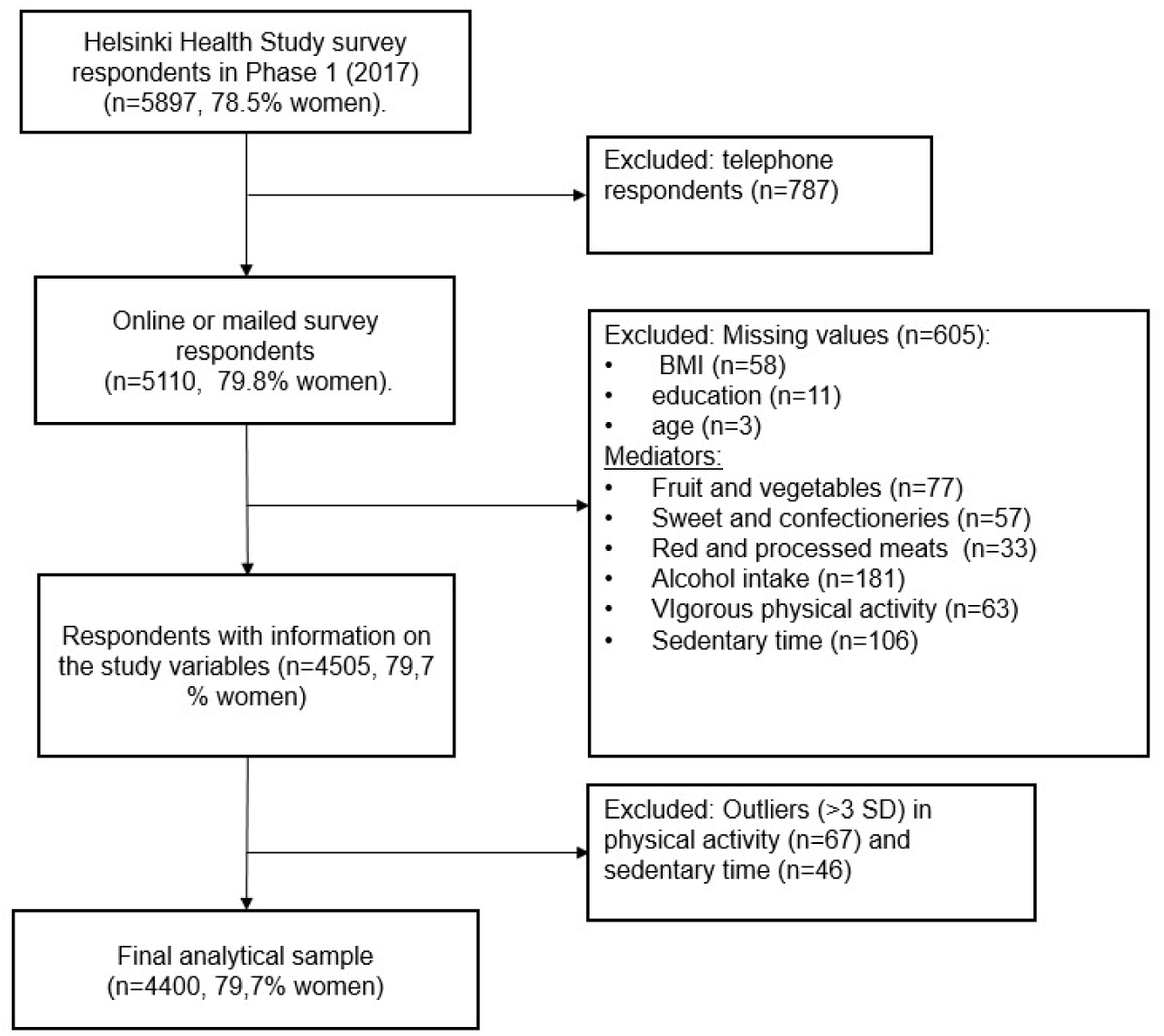
Flowchart of the exclusions leading to the final analytical sample. SD = Standard deviation

### Socioeconomic position and weight measures

Gender (male/female) was obtained from the surveys. SEP was measured using the highest educational attainment, categorized into three groups: upper secondary or vocational education or lower (low), bachelor’s degree (middle), and master’s degree or higher (high). BMI was computed from height and weight (kg/m²).

### Dietary factors and physical activity

We included six behaviors, four related to diet: fruit and vegetable consumption, red and processed meat consumption, sweets and confectionery consumption, alcohol consumption, and two related to physical activity during leisure time: VPA and sedentary time. These mediators were chosen because they are directly related to body weight through energy intake or energy expenditure.

Food consumption was assessed using a 14-item food frequency questionnaire (FFQ) covering the previous 4 weeks. Consumption of fruit and vegetables was calculated by summing four items: fresh vegetables, fresh fruits, cooked vegetables, and berries. Red and processed meat consumption was based on one questionnaire item, and sweets and confectionery consumption was calculated by summing two items, one on candy and chocolate and the other on coffee pastries and baked goods (biscuits/cookies, doughnuts, and other pastries). The variables were divided by four to obtain weekly servings. Alcohol consumption was assessed from participants’ reported frequency of consuming three beverage types: beer, cider, long drink and similar beverages; wine and other low- alcohol beverages; and spirits or other strong alcoholic beverages. Weekly alcohol consumption was calculated by summing the reported consumption frequencies across beverage types. One unit of alcohol refers to 12 grams of pure ethanol.

Participants reported their leisure-time and commuting physical activity during the past 12 months by classifying activities according to perceived intensity and estimating the average weekly duration at each intensity level. Intensity was converted to metabolic equivalents (METs) and multiplied by the duration of each intensity, following previous studies [29]. Only VPA (> 10 METs) was investigated as a mediator in the main analysis and total physical activity (sum of all the domains) in sensitivity analyses. Sedentary time was assessed as self-reported daily sitting time across four domains: at home while watching television or using a computer, at home while reading, during transport, and elsewhere. These were summed up and multiplied by seven to obtain time sitting per week. If a participant reported physical activity, sedentary time or alcohol consumption for some domain but left one domain blank, the missing value for that domain was set to zero, following prior studies [19,20].

### Statistical methods

Background characteristics of the participants were described by gender and SEP. Continuous variables were presented as means and categorical variables as counts and percentages. Age was categorized as 19–29 years and 30–39 years for descriptive analyses and treated as a continuous variable in mediation models, following previous studies [20]. Differences in dietary factors and physical activity across gender and SEP groups were examined descriptively and visualized using bar plots. The association between SEP and BMI was first examined visually and then tested using linear regression models. Interaction terms between SEP and gender were used to assess whether educational differences in BMI differed between women and men. Model assumptions, including the normality of residuals and heteroscedasticity, were assessed.

The mediating role of health dietary factors and physical activity in the association between SEP and BMI was examined using multiple parallel mediation models within a structural equation modeling framework in accordance with the conceptual model shown in Figure 2. All analyses were stratified by gender and adjusted for age. A three-category SEP variable (upper secondary or vocational education or lower/bachelor’s degree/master’s degree or higher) was created for men and women separately, with the highest education being the reference group. In each mediation model, SEP was specified as the exposure, BMI as the outcome, and dietary factors and physical activity as simultaneous mediators. All mediators were treated as continuous variables.

**Figure 2.**
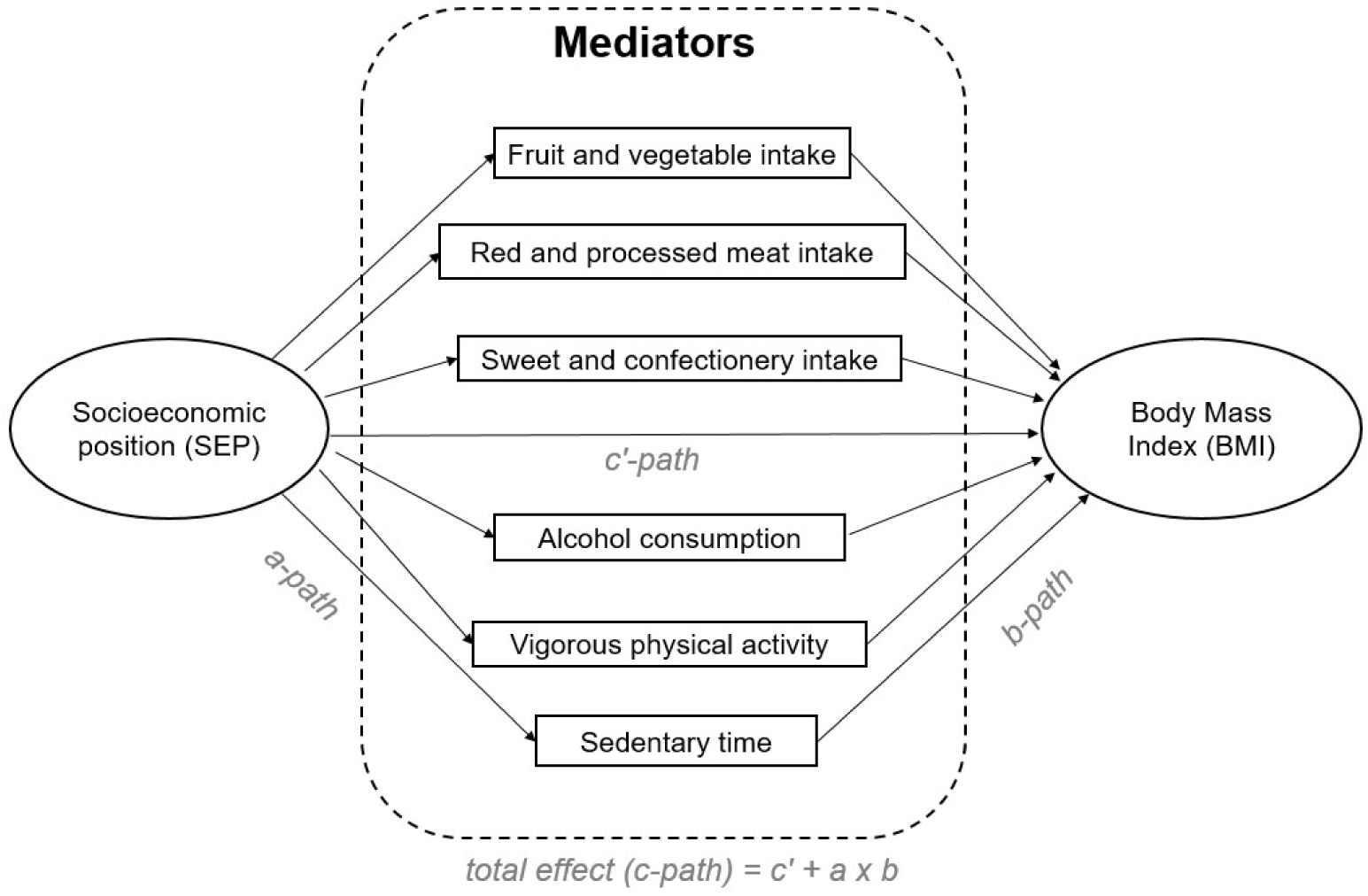
Conceptual mediation model of the association between socioeconomic position (SEP) and body mass index (BMI) through dietary factors and physical activity in the Helsinki Health Study cohort (n = 4,400) in Phase 1 (2017). The a-path represents the association between SEP and each mediator, the b-path represents the association between each mediator and BMI adjusted for SEP and the other mediators, the c-path represents the total association, and the c’-path represents the direct association after adjustment for mediators.

The mediation analyses were conducted using structural equation modeling and followed the standard path-analytic approach [30]. First, the total association (c-path) between SEP and BMI was estimated. Second, the associations between SEP and each mediator were estimated as a-paths.

Third, the associations between each mediator and BMI, adjusted for SEP and all other mediators, were estimated as b-paths. The direct effect (c’-path) represented the association between SEP and BMI after adjustment for all mediators. Mediator-specific indirect effects were calculated as the product of the corresponding a- and b-paths. The total indirect effect was calculated as the sum of all mediator-specific indirect effects. When the total effect (c-path) was statistically significant (p<0.05), the proportion mediated was calculated by dividing the indirect effect by the total effect. Indirect effects and confidence intervals were estimated using bootstrapped standard errors with 5,000 replications. Model parameters are presented as non-standardized regression coefficients. Non-standardized estimates were used to allow more direct comparison between women and men, as the focus was on gender differences rather than comparisons between mediators within genders.

Standardized estimates depend on variable variances, which may differ by gender. Results are reported separately for women and men as total effects, direct effects, total indirect effects and mediator-specific indirect effects. Models were fitted using the lavaan package in R (version 4.5.3, 2026-03-11). ChatGPT (OpenAI; accessed May 2025) was used to assist in generating R code for statistical analyses. All AI-generated code was reviewed, tested, and modified as necessary by the authors, who verified the accuracy and reproducibility of the results. A STROBE checklist is provided as a supplementary file (File S1).

### Additional follow-up analyses

To assess whether the observed mediation reflected socioeconomic differences in BMI already present at Phase 1, a secondary longitudinal analysis among those who had responded to the Phase 2 survey five years later in 2022 (n = 2581, 80% women) was conducted. The cross-sectional analysis was chosen as the main analysis to maximize statistical power, as 40% of participants were lost to follow-up between Phases 1 and 2. In the longitudinal model, Phase 2 BMI was used as the outcome variable, and the models were additionally adjusted for Phase 1 BMI. Education was taken from Phase 2 to consider new completed education between Phase 1 and 2.

## Results

### Descriptive statistics

As shown in Table 1, those with higher education were generally older than those in lower education groups. Mean BMI in the total sample was 25.2 kg/m² and increased with decreasing educational level in both women and men. Normal weight was most common in those with highest education, whereas overweight and obesity were more prevalent in the lower education groups, with overweight being more common among men.

**Table 1.** Descriptive characteristics of the Helsinki Health Study participants in Phase 1 (2017).

|  |  | **All** | **Women**<br>(n=3506, 79.7 %) |  |  | **Men**<br>(n=894, 20.3 %) |  |  | **P-value\*** |
|---|---|---|---|---|---|---|---|---|---|
|  |  |  | Master's degree or higher | Bachelor's degree | Upper secondary or vocational school or lower | Master's degree or higher | Bachelor's degree | Upper secondary or vocational school or lower |  |
| **Total** |  | 4400 | 1062 (24.1) | 1361 (30.1) | 1083 (24.6) | 257 (5.7) | 267 (6.1) | 370 (8.4) |  |
| **Age**<br>(n (%)) | 19–29 years | 1376 (31.3) | 178 (16.8) | 482 (35.4) | 475 (43.9) | 40 (15.6) | 69 (25.8) | 132 (35.7) | <0.001 |
|  | 30–39 years | 3022 (68.7) | 882 (83.2) | 879 (64.6) | 608 (56.1) | 217 (84.4) | 198 (74.2) | 238 (64.3) |  |
| **BMI, kg/m<sup>2</sup>**<br>(mean (SD)) |  | 25.24 (5.2) | 24.04 (4.5) | 24.72 (4.81) | 26.40 (6.55) | 25.20 (3.68) | 26.04 (3.74) | 26.63 (4.72) | <0.001 |
| **BMI (categorized)**<br>(n (%)) | Normal weight and under-weight | 2615 (59.4) | 756 (71.2) | 884 (65.0) | 557 (51.4) | 139 (54.1) | 120 (44.9) | 159 (43.0) | <0.001 |
|  | Overweight | 1147 (26.1) | 210 (19.8) | 302 (22.2) | 289 (26.7) | 96 (37.4) | 110 (41.2) | 140 (37.8) |  |
|  | Obesity | 638 (14.5) | 96 (9.0) | 175 (12.9) | 237 (21.9) | 22 (8.6) | 37 (13.9) | 71 (19.2) |  |

\* P-values were calculated using χ<sup>2</sup> test for categorical variables and a t- test for mean BMI
SD = standard deviation, BMI = body mass index

Figure 3 shows clear differences in dietary factors and physical activity across gender and SEP. Women reported a higher consumption fruit and vegetables than men in all education groups, and in both genders, consumption decreased with lower education. In contrast, red and processed meat consumption increased with decreasing education and was consistently higher among men than women. Sweets and confectionery consumption was also higher among women than men, whereas alcohol consumption was higher among men across all education groups, with little difference by education. VPA was higher among men than women and declined with decreasing education in both genders. Sedentary time showed the opposite educational pattern, tending to be higher in the lower education groups, particularly among men.

**Figure 3.**
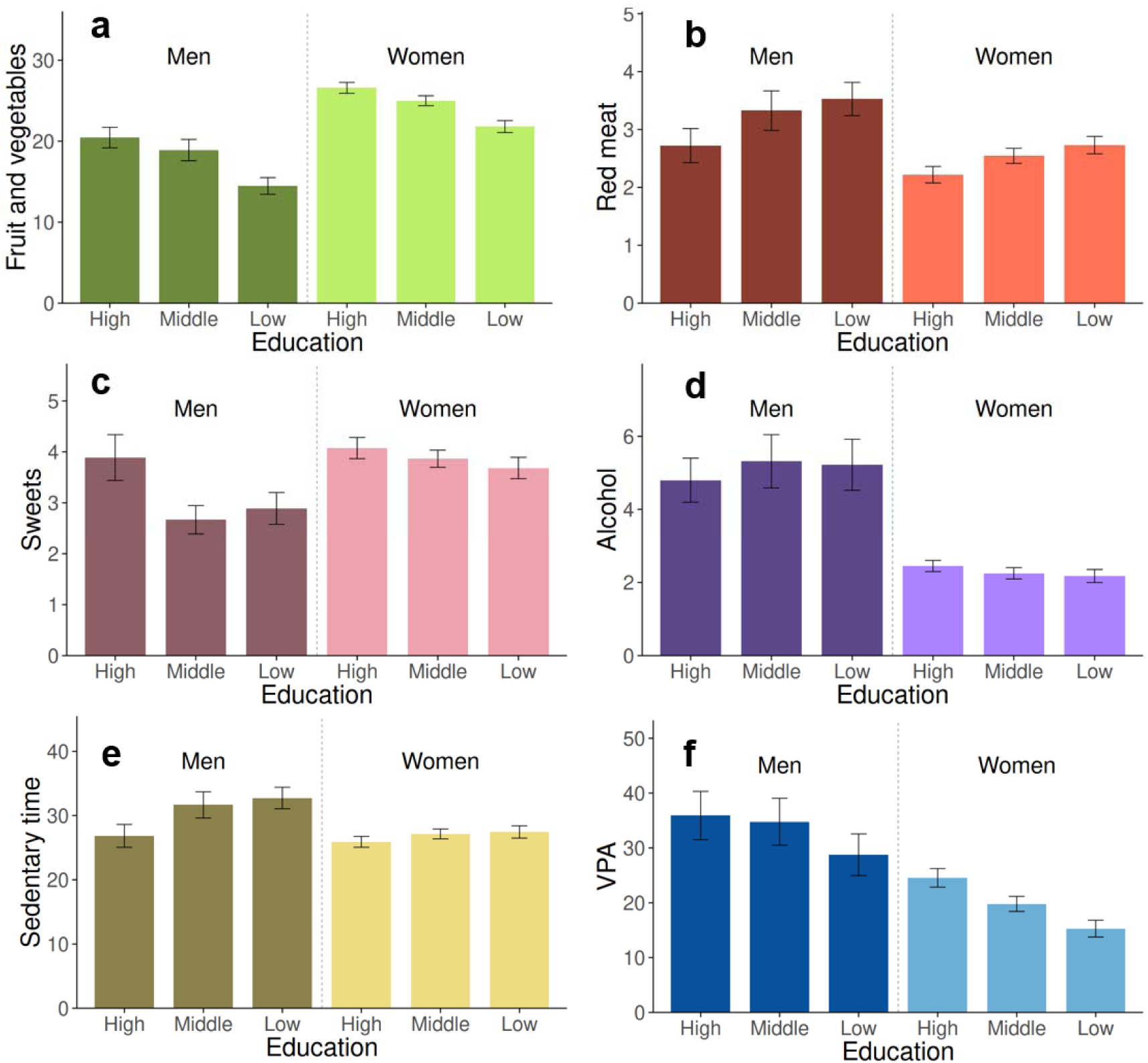
Differences in dietary factors and physical activity across gender and educational groups in the Helsinki Health Study cohort (n = 4,400) in Phase 1 (2017). Bars represent means and error bars represent their 95% confidence intervals. Panels show a) fruit and vegetable consumption (servings/week), b) red and processed meat consumption (servings/week), c) sweets and confectionery consumption (servings/week), d) total alcohol consumption from different beverage types (servings/week), e) sedentary time (hours/week), and f) vigorous physical activity (MET-hours/week). High education = master’s degree or higher, middle education = bachelor’s degree, low education = upper secondary or vocational school or lower, VPA = vigorous physical activity, MET = metabolic equivalent

Figure 4 shows an inverse association between SEP and BMI in both genders. Among men, the gradient was relatively modest, whereas among women it was steeper, although men had a higher mean BMI overall. Consequently, men had a higher BMI than women in the two higher education groups, but this gender difference narrowed in the lowest education group. Statistical testing showed that both educational level and gender were associated with BMI (p < 0.001), and the education × gender interaction was statistically significant (p = 0.037), suggesting a gender difference in the association between SEP and BMI.

**Figure 4.**
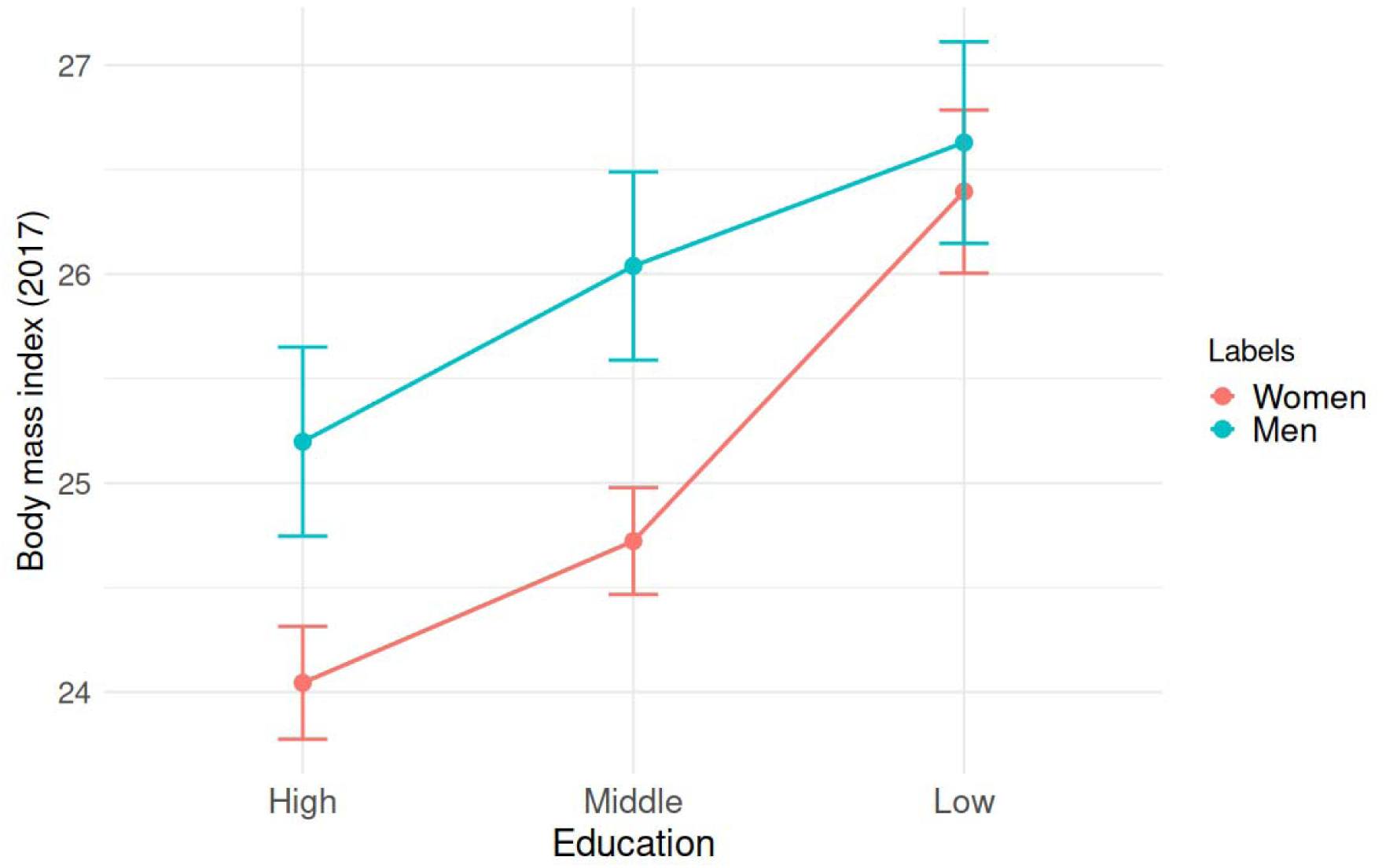
Gender differences in the association between socioeconomic position and body mass index in the Helsinki Health Study Cohort (n = 4,400, 80% women) in Phase 1 (2017). Points represent means and error bars represent their 95% confidence intervals. High education = master’s degree or higher, middle education = bachelor’s degree, low education = upper secondary or vocational school or lower, BMI = body mass index (kg/m²)

### Total effect of socioeconomic position on body mass index

Compared to participants with high education, the total effect of middle education on BMI was similar in women and men, at 0.98 (95% confidence interval (CI) 0.61, 1.37) and 0.97 (95% CI 0.33, 1.61), respectively. The association was stronger among participants with low education, particularly among women: compared with high education, the total effect of low education on BMI was 2.84 (95% CI 2.34, 3.35) in women and 1.67 (95% CI 0.99, 2.34) in men.

### Exposure–mediator relationship (a-path)

In both women and men, compared with those with high education, the middle and low education groups had significantly lower consumption of fruit and vegetables, higher consumption of red and processed meat, and lower VPA, as shown by the a-paths in Table 2. In addition, lower education was associated with lower alcohol consumption among women, and with higher sedentary time and lower sweets consumption among men. Lower education was also associated with lower sweets and confectionery consumption among women and lower alcohol consumption among men.

**Table 2.** The mediating role of dietary factors and physical activity in the age-adjusted education-BMI association among women and men.

|  |  |  | Association<br>between education and mediator | Association<br>between mediator and BMI* | Mediated effect (indirect effect) |  |
|---|---|---|---|---|---|---|
| **Mediator** | **Education**<br>(Ref: High education) | **Gender** | **a-path**<br>**(95% CI)** | **b-path**<br>**(95% CI)** | **a x b (95% CI)** | **Proportion mediated** |
| Fruit and vegetable consumption | Middle | W | -1.37 (-2.30, -0.45) | -0.03 (-0.05, -0.02) | 0.05 (0.01, 0.09) | 4.8% |
|  |  | M | -1.38 (-3.21, 0.43) | -0.01 (-0.04, 0.02) | 0.02 (-0.03, 0.08) | 1.7% |
|  | Low | W | -4.44 (-5.45, -3.40) | -0.03 (-0.05, -0.02) | 0.15 (0.08, 0.23) | 5.4% |
|  |  | M | -5.67 (-7.36, -3.99) | -0.01 (-0.04, 0.02) | 0.07 (-0.09, 0.23) | 4.0% |
| Red and processed meat consumption | Middle | W | 0.36 (0.16, 0.55) | 0.11 (0.04, 0.18) | 0.04 (0.01, 0.07) | 3.9% |
|  |  | M | 0.62 (0.16, 1.08) | 0.14 (0.04, 0.25) | 0.09 (0.01, 0.19) | 9.2% |
|  | Low | W | 0.56 (0.35, 0.78) | 0.11 (0.04, 0.18) | 0.06 (0.02, 0.11) | 2.1% |
|  |  | M | 0.83 (0.40, 1.25) | 0.14 (0.04, 0.25) | 0.12 (0.03, 0.23) | 7.2% |
| Sweets and confectionery consumption | Middle | W | -0.13 (-0.40, 0.15) | -0.03 (-0.08, 0.02) | 0.00 (-0.01, 0.02) | 0.3% |
|  |  | M | -1.16 (-1.71, -0.64) | -0.03 (-0.15, 0.09) | 0.04 (-0.12, 0.18) | 3.6% |
|  | Low | W | -0.26 (-0.56, 0.03) | -0.03 (-0.08, 0.02) | 0.01 (-0.01, 0.03) | 0.2% |
|  |  | M | -0.88 (-1.44, -0.32) | -0.03 (-0.15, 0.09) | 0.03 (-0.09, 0.15) | 1.6% |
| Alcohol consumption | Middle | W | -0.24 (-0.46, -0.02) | 0.04 (-0.01, 0.1) | -0.01 (-0.03, 0.00) | -1.1% |
|  |  | M | 0.51 (-0.41, 1.50) | 0.02 (-0.04, 0.09) | 0.01 (-0.03, 0.08) | 1.2% |
|  | Low | W | -0.33 (-0.58, -0.08) | 0.04 (-0.01, 0.1) | -0.02 (-0.04, 0.00) | -0.5% |
|  |  | M | 0.41 (-0.50, 1.38) | 0.02 (-0.04, 0.09) | 0.01 (-0.03, 0.07) | 0.5% |
| Vigorous physical activity | Middle | W | -5.77 (-7.99, -3.56) | -0.03 (-0.04, -0.03) | 0.18 (0.11, 0.26) | 18.6% |
|  |  | M | -1.46 (-7.83, 4.52) | -0.01 (-0.02, -0.01) | 0.02 (-0.06, 0.11) | 1.9% |
|  | Low | W | -10.89 (-13.19, -8.44) | -0.03 (-0.04, -0.03) | 0.35 (0.26, 0.44) | 12.1% |
|  |  | M | -7.78 (-13.71, -1.95) | -0.01 (-0.02, -0.01) | 0.10 (0.02, 0.21) | 5.8% |
| Sedentary time | Middle | W | 0.1 (-0.06, 0.26) | 0.22 (0.14, 0.30) | 0.02 (-0.01, 0.06) | 2.3% |
|  |  | M | 0.63 (0.24, 1.02) | 0.14 (0.01, 0.28) | 0.09 (0.01, 0.22) | 9.3% |
|  | Low | W | 0.1 (-0.09, 0.29) | 0.22 (0.14, 0.30) | 0.02 (-0.02, 0.07) | 0.8% |
|  |  | M | 0.74 (0.38, 1.1) | 0.14 (0.01, 0.28) | 0.11 (0.01, 0.24) | 6.3% |
| All mediators together | Middle | W | - | - | 0.28 (0.18, 0.39) | 29.0 % |
|  |  | M | - | - | 0.26 (0.02, 0.51) | 26.8 % |
|  | Low | W | - | - | 0.57 (0.44, 0.72) | 20.1 % |
|  |  | M | - | - | 0.42 (0.18, 0.68) | 25.3 % |

\*adjusted for socioeconomic position and other mediators
High education = master’s degree or higher, middle education = bachelor’s degree, low education = upper secondary or vocational school or lower, CI = confidence interval, W=women, M=men, BMI = body mass index

### Mediator–outcome relationship (b-path)

Higher consumption of fruit and vegetables was associated with lower BMI only among women, whereas higher VPA was associated with lower BMI in both genders, shown as the b-paths in Table 2. In contrast, higher red and processed meat consumption and greater sedentary time were associated with higher BMI among both women and men. The associations of VPA and sedentary time with BMI were stronger among women, whereas the association between red and processed meat and BMI was stronger among men. Sweets and confectionery and alcohol consumption were not significantly associated with BMI in either gender.

### Indirect effect of socioeconomic position on body mass index through dietary factors and physical activity

Among women, VPA showed the largest indirect effect, both for middle versus high education (0.18, 95% CI 0.11, 0.26; 18.6% mediated) and low versus high education (0.35, 95% CI 0.26, 0.44; 12.1% mediated). Smaller indirect effects were observed through consumption of fruit and vegetables among women (middle vs high: 0.05, 95% CI 0.01, 0.09; 4.8% mediated; low vs high: 0.15, 95% CI 0.08, 0.23; 5.4% mediated) and through consumption of red and processed meat in both women (middle vs high: 0.04, 95% CI 0.01, 0.07; 3.9% mediated; low vs high: 0.06, 95% CI 0.02, 0.11; 2.1% mediated) and men (middle vs high: 0.09, 95% CI 0.01, 0.19; 9.2% mediated; low vs high: 0.12, 95% CI 0.03, 0.23; 7.2% mediated). Among men, additional significant indirect pathways were observed through sedentary time (middle vs high: 0.09, 95% CI 0.01, 0.22; 9.3% mediated; low vs high: 0.11, 95% CI 0.01, 0.24; 6.3% mediated) and, for the low versus high education comparison, through VPA (0.10, 95% CI 0.02, 0.21; 5.3% mediated). Red and processed meat consumption showed larger absolute and relative indirect effects in men than in women, whereas no clear indirect effects were observed for sweets and confectionery consumption or alcohol consumption in either gender. Taken together, all mediators explained 29.0% of the education–BMI association among women in the middle versus high education comparison and 20.1% in the low versus high comparison; among men, the corresponding proportions were 26.8% and 25.3%, respectively.

### Additional follow-up analysis with adjustment for baseline body mass index

After excluding those who had not responded to the follow-up survey, the number of participants was 2581. The sex distribution remained almost unchanged, but the follow-up sample was somewhat older and more highly educated. The socioeconomic gradient in BMI remained clearly visible in Phase 2 (Figure S1). Among women, mean BMI increased by 1.1 kg/m² in the high education group, 1.3 kg/m² in the middle education group, and 1.3 kg/m² in the low education group, reaching 25.4 kg/m², 26.0 kg/m², and 28.5 kg/m² in 2022, respectively. Among men, mean BMI increased by 0.7 kg/m² in the high education group, 0.9 kg/m² in the middle education group, and 0.9 kg/m² in the low education group, reaching 26.0 kg/m², 27.3 kg/m², and 28.0 kg/m² in 2022, respectively. Thus, BMI increased over follow-up in all education groups, but the highest BMI levels were consistently observed in the low education group, and the increase was more pronounced among women. The categorical BMI distribution showed a similar development, with the proportion of overweight and obesity increasing and normal weight decreasing. The complete characteristics of the follow-up sample are presented in Table S1.

The gender interaction in the SEP-BMI association in Phases 1 and 2 among those who had responded to the follow-up survey (n=2581) is shown in Figure S1. By 2022, mean BMI had increased in all education groups, but the inverse gradient remained. The gender differences had narrowed, meaning that weight gain was more pronounced in women. At follow-up, education remained strongly associated with BMI (p < 0.001), whereas gender no longer reached statistical significance (p = 0.085). However, the education × gender interaction remained significant (p = 0.040), meaning that the gender difference in the association between SEP and BMI remained.

After accounting for baseline BMI in the follow-up analysis, the total effects of SEP on Phase 2 BMI were small and not statistically significant in any group, as is shown in Table S2. The a-paths showed a broadly similar pattern with lower educational level being still associated with lower consumption of fruit and vegetables and higher red and processed meat consumption. Lower sweets and confectionery consumption and more sedentary time were associated with lower education in men, whereas lower alcohol consumption and less VPA were associated with lower education in women. However, the b-paths linking the mediators to follow-up BMI were clearly weaker than in the cross-sectional model with only red and processed meat consumption remaining positively associated with follow-up BMI among women and fruit and vegetable and sweets and confectionery consumption in men.

Accordingly, the indirect effects were small and inconsistent. Among women, there was some evidence of mediation through red and processed meat consumption and through VPA. Among men, the significant indirect effects were observed for consumption of fruit and vegetables and sweets and confectioneries. The summed indirect effects across all mediators were not statistically significant in any group. Since the total effects of SEP on Phase 2 BMI were not statistically significant, the mediated proportions were not calculated. The longitudinal model adjusting for baseline BMI suggests that the educational differences in BMI were present already at Phase 1 and that Phase 2 BMI was largely explained by prior BMI rather than by subsequent BMI change.

## Discussion

### Main findings

Our main finding was that the association between SEP and BMI was stronger among women than among men, and that this difference was partly explained by dietary factors and physical activity. Consumption of red and processed meat was an important mediator in both genders, although it explained a larger part of the SEP–BMI association among men. VPA was the most prominent mediator among women, followed by consumption of fruit and vegetables. Sedentary time was a significant mediator only among men, while alcohol and sweets and confectionery consumption did not mediate the SEP-BMI association in either gender. Additional analyses with 5-year follow-up BMI adjusted for baseline BMI suggested that educational differences in follow-up BMI were largely explained by baseline BMI instead of the mediators.

### Comparison to previous literature

The steeper SEP–BMI gradient in women than in men has been well established in previous literature, both in the Finnish population and internationally [8,11]. SEP tends to be associated with obesity regardless of the indicator used, but in studies examining multiple SEP indicators, the associations with body weight have generally been more consistent among women and more variable among men [31–34]. Because we used education as the SEP indicator in this study, it remains to be seen whether the observed gender differences in SEP-BMI association would be equally consistent if other SEP indicators were used.

Lower SEP was associated with a less favorable behavioral profile in both genders, including lower consumption of fruit and vegetables, higher consumption of red and processed meat, and lower VPA. The educational differences in diet quality were in line with the Finnish national dietary surveys [12] as well as international evidence, where the clearest support is seen for fruit and vegetable consumption [13,35]. Regarding physical activity and sedentary time, we observed clear socioeconomic gradients in VPA, which is consistent with previous literature [22,36]. Higher levels of VPA among the highly educated may reflect greater participation in organized sports. The pattern for sedentary time was less clear, although the socioeconomic gradient appeared somewhat stronger among men. These inconsistencies may be explained by differences between domains [20]. Because our study measured only leisure-time vigorous activity, the observed socioeconomic patterning may reflect the possibility that lower educated individuals accumulate more occupational physical activity and less occupational sedentary time, whereas higher educated individuals engage more in leisure-time physical activity but may also have more sedentary work. Additionally, gender differences related to dietary factors and physical activity were consistent with prior national and international evidence [12,37,38].

There are some studies examining gender differences in the mediators of the SEP–BMI association using similar methodology to ours. In a Finnish study consumption of fruit and vegetables, self-rated dietary quality and sedentary time mediated the association in both genders, while leisure-time physical activity and alcohol were mediators only in women and smoking only in men [39]. Another study found significant indirect effects through consumption of fruit and vegetables only among women [40]. Lower levels of physical activity and a higher consumption of unhealthy savory foods were important mediators in an Australian cross-sectional study, with alcohol consumption and diet soft drink intake being additional mediators among women [41]. In the only longitudinal study investigating the mediators of the SEP-BMI association, conducted in the previously mentioned cohort, the association was mediated by physical activity, intake of nutrient-rich foods, and diet soft drink consumption, with the mediation effects being clearly stronger among women [42]. In our study, consumption of fruit and vegetables and VPA were statistically significant mediators among women, in line with the earlier findings [40,42]. However, alcohol consumption was not a statistically significant mediator in either gender, and sedentary time was found to mediate the associations only among men. Despite the partly different behaviors included across studies, the overall magnitudes of the individual mediators explaining the SEP–BMI association were broadly similar to our results [39–42].

We also tested subjective sleep quality and total physical activity as additional mediators. These variables did not show clear or statistically significant indirect effects and were therefore not included in the main mediation models. Because not all mediators were normally distributed, we also examined median values in the descriptive analysis. For all mediators except meat, socioeconomic gradients and gender differences were consistent with the mean-based results. For red and processed meat consumption, the differences disappeared, with a median intake of 3 servings per week in all groups. This suggests that the distribution between groups differs, which was confirmed visually using histograms, although the differences were quite small (Figure S2). Highly educated women and highly educated men had relatively more observations in the lower range of consumption, whereas women and men with low education had relatively more observations in the higher range.

In our follow-up analysis, BMI increased in both genders and in all educational groups during the 5-year follow-up, which is in line with other Finnish studies [3]. However, we observed a faster increase in BMI among women than among men. The gender differences and educational gradients remained still visible, and previous research suggests that socioeconomic differences in body weight are relatively stable over time [43]. In additional mediation analyses with a 5-year follow-up BMI adjusted for baseline BMI, educational differences in BMI were largely explained by baseline BMI rather than by the mediators. In one longitudinal study, BMI gain was partly explained by health behaviors [42]. However, that study had a 13-year follow-up, during which educational differences in BMI gain were detected. It is therefore possible that our 5-year follow-up was too short to detect clear differences in BMI change. BMI increased rather similarly across educational groups, so adjusting for baseline BMI largely attenuated the educational differences.

### Methodological considerations

Regarding the study population, this study used a large occupational cohort of young adults aged 19– 39 years, including both manual and non-manual employees. However, the sample was mostly women (80%), which likely reduced statistical power in the male subgroup and may partly explain why some associations among men were not statistically significant. Although the data may be generalized with caution to the working-age Finnish population, they cannot be directly generalized to the entire adult population because the cohort consisted only of employees of the City of Helsinki. Thus, a healthy worker effect is likely, and the lowest SEP group in this study does not represent the most socioeconomically disadvantaged groups in society, such as the long-term unemployed [44].

We also excluded telephone interviewees, and our prior studies have shown that those who responded via email or mail are more often women and more highly educated [27]. Due to missing values, we conducted a sensitivity analysis using multiple imputation by chained equations (MICE)[45]. The results were highly consistent with the complete-case analyses: the effect sizes were similar, while the confidence intervals were narrower, reflecting the larger analytic sample (Table S3).

Regarding measurements, all variables were based on self-report. Self-reported height and weight may introduce bias, although they are generally considered sufficiently accurate for epidemiological research [46,47]. Additionally, both predictors and outcomes were measured by questionnaire, so common method bias is possible [48]. The dietary measure was based on a short FFQ that was not formally validated and did not include portion sizes, meaning that absolute consumption could not be estimated. However, short FFQs have been shown to be suitable for assessing group-level differences in frequently consumed foods [49,50] and portion-size questions may contribute relatively little to variance in intake [51]. The similarity between our results and the national dietary survey, which uses a more precise data collection method, supports our FFQ’s validity [12]. Similarly, the physical activity and sedentary time questionnaires were not validated in this cohort, although comparable measures have shown acceptable validity in epidemiological studies [52,53].

Regarding statistical analysis, our cross-sectional design does not allow for causal conclusions. We conducted a follow-up analysis, but it did not completely overcome this limitation because dietary factors and physical activity were measured at the same time as SEP. Additionally, the follow-up sample was smaller and more selective. We also adjusted the mediation models only for age, so some residual confounding may remain. However, the statistical framework of structural equation is built on the previous findings suggesting a causal relationship between SEP and BMI through health behaviors.

### Implications and significance

Our results suggest that socioeconomic differences in BMI are not explained by a single behavioral pathway, but rather by a myriad of factors, with some mechanisms being gender specific. For example, improving VPA may be relevant among low-SEP women, whereas reducing sedentary time may be more relevant among low-SEP men. At the same time, consumption of red and processed meat emerged as a common pathway in both genders. The stronger SEP–BMI gradient observed among women suggests that women in lower SEP may be more vulnerable to weight gain, so health policies should focus on this group.

Because these six mediators explained only about one quarter of the SEP–BMI association, the findings indicate the importance of other mechanisms, including psychosocial stress, neighborhood environment, working conditions and access to healthy food and exercise opportunities being potential mediators of the SEP-BMI association. Future research is needed to identify additional mediators of the SEP–BMI association to better understand why the association is more pronounced in women. Future studies would also benefit from repeated measurements of both mediators and BMI, so that changes over time and causal pathways could be examined more clearly.

## Conclusions

Taken together, our findings suggest that different behavioral mechanisms may underlie socioeconomic differences in BMI in men and women. VPA and fruit and vegetable consumption were significant mediators of the SEP-BMI association in women and sedentary time in men, while red and processed meat intake acted as a mediator in both genders. The association between SEP and BMI was stronger among women than among men, and the 5-year increase in BMI was more pronounced in women, especially among those with the lowest education. These results suggest that prevention of weight gain and the reduction of SEP-related health inequalities should focus on gender-specific approaches. At the same time, because only about one quarter of the SEP–BMI association was explained by the included dietary factors and physical activity patterns, broader structural, social, and environmental factors are likely to play an important role in maintaining weight-related inequalities.

## Author Contributions

Emma Järvensivu: data analysis, methodology, writing (original draft, and final manuscript), review and editing; Tea Lallukka: methodology, supervision, validation, and writing (review and editing); Lari Järvensivu: data analysis, methodology, validation, and writing (review and editing); Ossi Rahkonen: funding acquisition, methodology, validation, and writing (review and editing); Morris Viitanen: supervision, validation, and writing (review and editing). All authors reviewed and approved the final version of the manuscript.

## Acknowledgements

We thank the City of Helsinki, Finland, and all participating employees.

We acknowledge the use of ChatGPT (OpenAI; accessed May 2025) to assist with the drafting of the abstract and to suggest improvements to the organization of the Discussion and Introduction sections. All AI-generated suggestions were critically reviewed, revised and approved by the authors, who take full responsibility for the accuracy and integrity of the work.

## Conflict of Interest

The authors have no conflicts of interest to declare.

## Funding Statement

Authors Emma Järvensivu, Lari Järvensivu and Ossi Rahkonen are supported by the Ministry of Education and Culture, Finland [grant number: #OKM/76/626/2023]. Ossi Rahkonen is supported by the Juho Vainio Foundation [grant number: #202500047]. The funders had no role in the study design; data collection, management, analysis, or interpretation; manuscript conception, planning, or writing; or the decision to submit the manuscript for publication.

## Statement of Ethics

The Helsinki Health Study’s research plan received a favorable statement from the ethical board of the Faculty of Medicine at the University of Helsinki in February 2017 (decision nr. 01/2017) and an updated approval by the City of Helsinki in April 2017 (decision nr. HEL 2017-004490). Throughout all phases of the study, ethical standards consistent with the 1964 Helsinki Declaration were upheld.

Participants were fully informed about the study’s objectives, the voluntary nature of their participation, data confidentiality and handling procedures. Written informed consent was obtained from participants prior to the study. No compensation was offered to participants for their involvement in the study. A few gift certificates of small monetary value (€50) were drawn among all respondents.

## Data Availability

The Helsinki Health Study survey data cannot be made publicly available due to strict data protection laws and regulations. The data can only be used for scientific research. More information on the survey data can be requested from the Helsinki Health Study research group.

## Notes

### Competing Interest Statement

The authors have declared no competing interest.

### Author Declarations

The ethical board of the Faculty of Medicine at the University of Helsinki gave ethical approval in February 2017 (decision nr. 01/2017) The City of Helsinki in April 2017 (decision nr. HEL 2017-004490) gave approval for this work. Throughout all phases of the study, ethical standards consistent with the 1964 Helsinki Declaration were upheld.

## References

1 Kagenaar E, Van Hemelrijck WMJ, Kunst AE, Janssen F. Long-Term Trends in Obesity Prevalence by Socio-Economic Group in Five European Countries and the USA: The Relevance of the Diffusion of Innovations Theory. Obes Facts. 2022;15(6):753–61. doi:10.1159/000527070

2 Obesity and overweight [Internet]. [cited 2026 May 27]. Available from: https://www.who.int/news-room/fact-sheets/detail/obesity-and-overweight

3 Lundqvist A, Jääskeläinen T, Lehtoranta L, Aspholm S, Vessari H, Ojanen A, et al. Age-specific changes in obesity and associated cardiometabolic risk factors: a two-decade study of the Finnish adults. Eur J Public Health. 2026 Feb 1;36(1):162–8. doi:10.1093/eurpub/ckaf186

4 NCD Risk Factor Collaboration (NCD-RisC), Zhou B, Phelps NH, Galeazzi A, O’Driscoll ON, Bennett JE, et al. Obesity rise plateaus in developed nations and accelerates in developing nations. Nature. 2026 May 14;653(8114):510–8. doi:10.1038/s41586-026-10383-0

5 Galobardes B, Lynch J, Smith GD. Measuring socioeconomic position in health research. Br Med Bull. 2007 Feb 6;81–82(1):21–37. doi:10.1093/bmb/ldm001

6 Braveman PA, Cubbin C, Egerter S, Chideya S, Marchi KS, Metzler M, et al. Socioeconomic Status in Health Research: One Size Does Not Fit All. JAMA. 2005 Dec 14;294(22):2879. doi:10.1001/jama.294.22.2879

7 Newton S, Braithwaite D, Akinyemiju TF. Socio-economic status over the life course and obesity: Systematic review and meta-analysis. PLOS ONE. 2017 May 16;12(5):e0177151. doi:10.1371/journal.pone.0177151

8 Witkam R, Gwinnutt JM, Humphreys J, Gandrup J, Cooper R, Verstappen SMM. Do associations between education and obesity vary depending on the measure of obesity used? A systematic literature review and meta-analysis. SSM - Popul Health. 2021 Sep;15:100884. doi:10.1016/j.ssmph.2021.100884

9 Autret K, Bekelman TA. Socioeconomic Status and Obesity. J Endocr Soc. 2024 Sep 26;8(11):bvae176. doi:10.1210/jendso/bvae176

10 Lahti-Koski M. Body mass index and obesity among adults in Finland: Trends and determinants [Doctoral Dissertation] [Internet]. [Helsinki]: University of Helsinki; 2001. Available from: http://hdl.handle.net/10138/20332

11 Prättälä R, Sippola R, Lahti-Koski M, Laaksonen MT, Mäkinen T, Roos E. Twenty-five year trends in body mass index by education and income in Finland. BMC Public Health. 2012 Dec;12(1):936. doi:10.1186/1471-2458-12-936

12 Ravitsemus Suomessa: FinRavinto 2017 -tutkimus [Internet]. THL; 2018 [cited 2026 May 26]. Available from: https://www.julkari.fi/handle/11111/31994

13 De Irala-Estévez J, Groth M, Johansson L, Oltersdorf U, Prättälä R, Martínez-González M. A systematic review of socio-economic differences in food habits in Europe: consumption of fruit and vegetables. Eur J Clin Nutr. 2000 Sep 1;54(9):706–14. doi:10.1038/sj.ejcn.1601080

14 Kristoffersen E, Hjort SL, Thomassen LM, Arjmand EJ, Perillo M, Balakrishna R, et al. Umbrella Review of Systematic Reviews and Meta-Analyses on the Consumption of Different Food Groups and the Risk of Overweight and Obesity. Nutrients. 2025 Feb 13;17(4):662. doi:10.3390/nu17040662

15 Händel MN, Rohde JF, Larsen SC, Sørensen TIA, Heitmann BL. Processed Meat Intake and Changes in Weight, Waist, Body Mass Index, and Fat Mass: Systematic Literature Review, Meta-Analysis, and GRADE Assessment of Cohort Studies of Adults. Nutr Rev. 2025 Oct 1;83(10):1827–42. doi:10.1093/nutrit/nuaf030

16 Schlesinger S, Neuenschwander M, Schwedhelm C, Hoffmann G, Bechthold A, Boeing H, et al. Food Groups and Risk of Overweight, Obesity, and Weight Gain: A Systematic Review and Dose-Response Meta-Analysis of Prospective Studies. Adv Nutr. 2019 Mar;10(2):205–18. doi:10.1093/advances/nmy092

17 Sayon-Orea C, Martinez-Gonzalez MA, Bes-Rastrollo M. Alcohol consumption and body weight: a systematic review. Nutr Rev. 2011 Aug;69(8):419–31. doi:10.1111/j.1753-4887.2011.00403

18 Jakicic JM, Powell KE, Campbell WW, Dipietro L, Pate RR, Pescatello LS, et al. Physical Activity and the Prevention of Weight Gain in Adults: A Systematic Review. Med Sci Sports Exerc. 2019 Jun;51(6):1262–9. doi:10.1249/MSS.0000000000001938

19 Päivärinne V, Salmela J, Lahti J, Pietiläinen O, Karjala A, Kouvonen A, et al. Life-course socioeconomic circumstances and changes in leisure-time physical activity among young and early midlife employees. Int J Equity Health. 2026 Jan 30;25(1):54. doi:10.1186/s12939-026-02769-3

20 Kekäläinen E, Rahkonen O, Päivärinne V, Nurminen H, Lallukka T. Childhood and current socioeconomic position as determinants of sedentary time among young and early midlife employees. Eur J Public Health. 2025 Oct 1;35(5):916–24. doi:10.1093/eurpub/ckaf152

21 Beaulieu K, Hopkins M, Blundell J, Finlayson G. Homeostatic and non-homeostatic appetite control along the spectrum of physical activity levels: An updated perspective. Physiol Behav. 2018 Aug;192:23–9. doi:10.1016/j.physbeh.2017.12.032

22 Beenackers MA, Kamphuis CB, Giskes K, Brug J, Kunst AE, Burdorf A, et al. Socioeconomic inequalities in occupational, leisure-time, and transport related physical activity among European adults: A systematic review. Int J Behav Nutr Phys Act. 2012;9(1):116. doi:10.1186/1479-5868-9-116

23 Silveira EA, Mendonça CR, Delpino FM, Elias Souza GV, Pereira De Souza Rosa L, De Oliveira C, et al. Sedentary behavior, physical inactivity, abdominal obesity and obesity in adults and older adults: A systematic review and meta-analysis. Clin Nutr ESPEN. 2022 Aug;50:63–73. doi:10.1016/j.clnesp.2022.06.001

24 Biddle SJH, Bengoechea García E, Pedisic Z, Bennie J, Vergeer I, Wiesner G. Screen Time, Other Sedentary Behaviours, and Obesity Risk in Adults: A Review of Reviews. Curr Obes Rep. 2017 Jun;6(2):134–47. doi:10.1007/s13679-017-0256-9

25 Curran F, Davis ME, Murphy K, Tersigni N, King A, Ngo N, et al. Correlates of physical activity and sedentary behavior in adults living with overweight and obesity: A systematic review. Obes Rev. 2023 Nov;24(11):e13615. doi:10.1111/obr.13615

26 Rodriguez Roca B, Tully MA, Sansano-Nadal O, Caserotti P, Coll-Planas L, Roqué M, et al. Is education level, as a proxy for socio-economic position, related to device-measured and self-reported sedentary behavior in European older adults? A cross-sectional study from the SITLESS project. Front Public Health. 2023 Dec 19;11:1296821. doi:10.3389/fpubh.2023.1296821

27 Lallukka T, Pietiläinen O, Jäppinen S, Laaksonen M, Lahti J, Rahkonen O. Factors associated with health survey response among young employees: a register-based study using online, mailed and telephone interview data collection methods. BMC Public Health. 2020 Dec;20(1):184. doi:10.1186/s12889-020-8241-8

28 City of Helsinki. Staff Report 2022 [Internet]. Helsinki; 2022. Available from: https://www.hel.fi/static/kanslia/julkaisut/2023/city-of-helsinki-staff-report-2022.pdf

29 Kujala UM, Kaprio J, Sarna S, Koskenvuo M. Relationship of Leisure-Time Physical Activity and Mortality: The Finnish Twin Cohort. JAMA. 1998 Feb 11;279(6):440. doi:10.1001/jama.279.6.440

30 MacKinnon DP, Fairchild AJ, Fritz MS. Mediation Analysis. Annu Rev Psychol. 2007 Jan 1;58(1):593–614. doi:10.1146/annurev.psych.58.110405.085542

31 Laaksonen M, Sarlio-Lähteenkorva S, Lahelma E. Multiple Dimensions of Socioeconomic Position and Obesity among Employees: The Helsinki Health Study. Obes Res. 2004 Nov;12(11):1851–8. doi:10.1038/oby.2004.230

32 Loman T, Lallukka T, Laaksonen M, Rahkonen O, Lahelma E. Multiple socioeconomic determinants of weight gain: the Helsinki Health Study. BMC Public Health. 2013 Dec;13(1):259. doi:10.1186/1471-2458-13-259

33 Wardle J, Waller J, Jarvis MJ. Sex Differences in the Association of Socioeconomic Status With Obesity. Am J Public Health. 2002 Aug;92(8):1299–304. doi:10.2105/AJPH.92.8.1299

34 Ogden CL, Fakhouri TH, Carroll MD, Hales CM, Fryar CD, Li X, et al. Prevalence of Obesity Among Adults, by Household Income and Education — United States, 2011–2014. MMWR Morb Mortal Wkly Rep. 2017 Dec 22;66(50):1369–73. doi:10.15585/mmwr.mm6650a1

35 Carrillo-Alvarez E, Rifà-Ros R, Salinas-Roca B, Costa-Tutusaus L, Lamas M, Rodriguez-Monforte M. Diet-Related Health Inequalities in High-Income Countries: A Scoping Review of Observational Studies. Adv Nutr. 2025 Jun;16(6):100439. doi:10.1016/j.advnut.2025.100439

36 O’Donoghue G, Kennedy A, Puggina A, Aleksovska K, Buck C, Burns C, et al. Socio-economic determinants of physical activity across the life course: A “DEterminants of DIet and Physical ACtivity” (DEDIPAC) umbrella literature review. PLOS ONE. 2018 Jan 19;13(1):e0190737. doi:10.1371/journal.pone.0190737

37 Strain T, Flaxman S, Guthold R, Semenova E, Cowan M, Riley LM, et al. National, regional, and global trends in insufficient physical activity among adults from 2000 to 2022: a pooled analysis of 507 population-based surveys with 5·7 million participants. Lancet Glob Health. 2024 Aug;12(8):e1232–43. doi:10.1016/S2214-109X(24)00150-5

38 Oja S, Hakamäki M, Syväoja H, Lehtonen K. Equality in sports and physical activity in Finland in 2025 [Internet]. Helsinki: Ministry of Education and Culture; 2026 [cited 2026 May 27]. Available from: https://julkaisut.valtioneuvosto.fi/handle/11111/13433

39 Borodulin K, Zimmer C, Sippola R, Mäkinen TE, Laatikainen T, Prättälä R. Health Behaviours as Mediating Pathways between Socioeconomic Position and Body Mass Index. Int J Behav Med. 2012 Mar;19(1):14–22. doi:10.1007/s12529-010-9138-1

40 Ward H, Tarasuk V, Mendelson R, McKeown-Eyssen G. An exploration of socioeconomic variation in lifestyle factors and adiposity in the Ontario Food Survey through structural equation modeling. Int J Behav Nutr Phys Act. 2007 Dec;4(1):8. doi:10.1186/1479-5868-4-8

41 Gearon E, Backholer K, Hodge A, Peeters A. The mediating role of dietary factors and leisure time physical activity on socioeconomic inequalities in body mass index among Australian adults. BMC Public Health. 2013 Dec;13(1):1214. doi:10.1186/1471-2458-13-1214

42 Gearon E, Peeters A, Ng W, Hodge A, Backholer K. Diet and physical activity as possible mediators of the association between educational attainment and body mass index gain among Australian adults. Int J Public Health. 2018 Sep;63(7):883–93. doi:10.1007/s00038-018-1100-z

43 Cleland V, Tian J, Buscot MJ, Magnussen CG, Bazzano L, Burns TL, et al. Body-mass index trajectories from childhood to mid-adulthood and their sociodemographic predictors: Evidence from the International Childhood Cardiovascular Cohort (i3C) Consortium. eClinicalMedicine. 2022 Jun;48:101440. doi:10.1016/j.eclinm.2022.101440

44 Li CY, Sung FC. A review of the healthy worker effect in occupational epidemiology. Occup Med. 1999;49(4):225–9. doi:10.1093/occmed/49.4.225

45 Azur MJ, Stuart EA, Frangakis C, Leaf PJ. Multiple imputation by chained equations: what is it and how does it work? Int J Methods Psychiatr Res. 2011 Mar;20(1):40–9. doi:10.1002/mpr.329

46 Davies A, Wellard-Cole L, Rangan A, Allman-Farinelli M. Validity of self-reported weight and height for BMI classification: A cross-sectional study among young adults. Nutrition. 2020 Mar;71:110622. doi:10.1016/j.nut.2019.110622

47 Stommel M, Schoenborn CA. Accuracy and usefulness of BMI measures based on self-reported weight and height: findings from the NHANES & NHIS 2001-2006. BMC Public Health. 2009 Dec;9(1):421. doi:10.1186/1471-2458-9-421

48 Podsakoff PM, MacKenzie SB, Lee JY, Podsakoff NP. Common method biases in behavioral research: A critical review of the literature and recommended remedies. J Appl Psychol. 2003;88(5):879–903. doi:10.1037/0021-9010.88.5.879

49 Osler M, Heitmann BL. The Validity of a Short Food Frequency Questionnaire and its Ability to Measure Changes in Food Intake: A Longitudinal Study. Int J Epidemiol. 1996;25(5):1023–9. doi:10.1093/ije/25.5.1023

50 Lillegaard ITL, Øverby NC, Andersen LF. Evaluation of a short food frequency questionnaire used among Norwegian children. Food Nutr Res. 2012 Jan;56:6399. doi:10.3402/fnr.v56i0.6399

51 Noethlings U, Hoffmann K, Bergmann MM, Boeing H. Portion Size Adds Limited Information on Variance in Food Intake of Participants in the EPIC-Potsdam Study. J Nutr. 2003 Feb;133(2):510–5. doi:10.1093/jn/133.2.510

52 Waller K, Kaprio J, Kujala UM. Associations between long-term physical activity, waist circumference and weight gain: a 30-year longitudinal twin study. Int J Obes. 2008 Feb;32(2):353–61. doi:10.1038/sj.ijo.0803692

53 Marshall AL, Miller YD, Burton NW, Brown WJ. Measuring Total and Domain-Specific Sitting: A Study of Reliability and Validity. Med Sci Sports Exerc. 2010 Jun;42(6):1094–102. doi:10.1249/MSS.0b013e3181c5ec18

