## Supplementary figures and images for "Gender Differences in Socioeconomic Inequalities in Body Mass Index and Health-Related Mediators Among Young and Early Midlife Finns"

### Figure S1

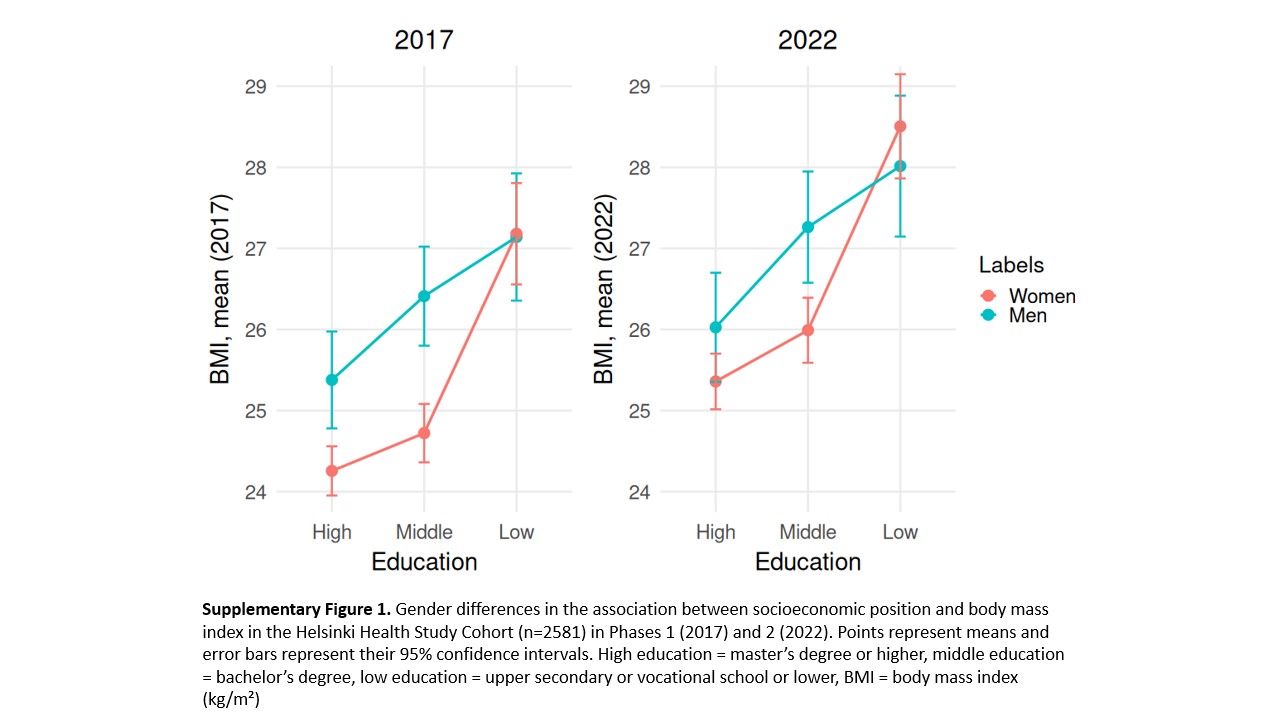

### Figure S2

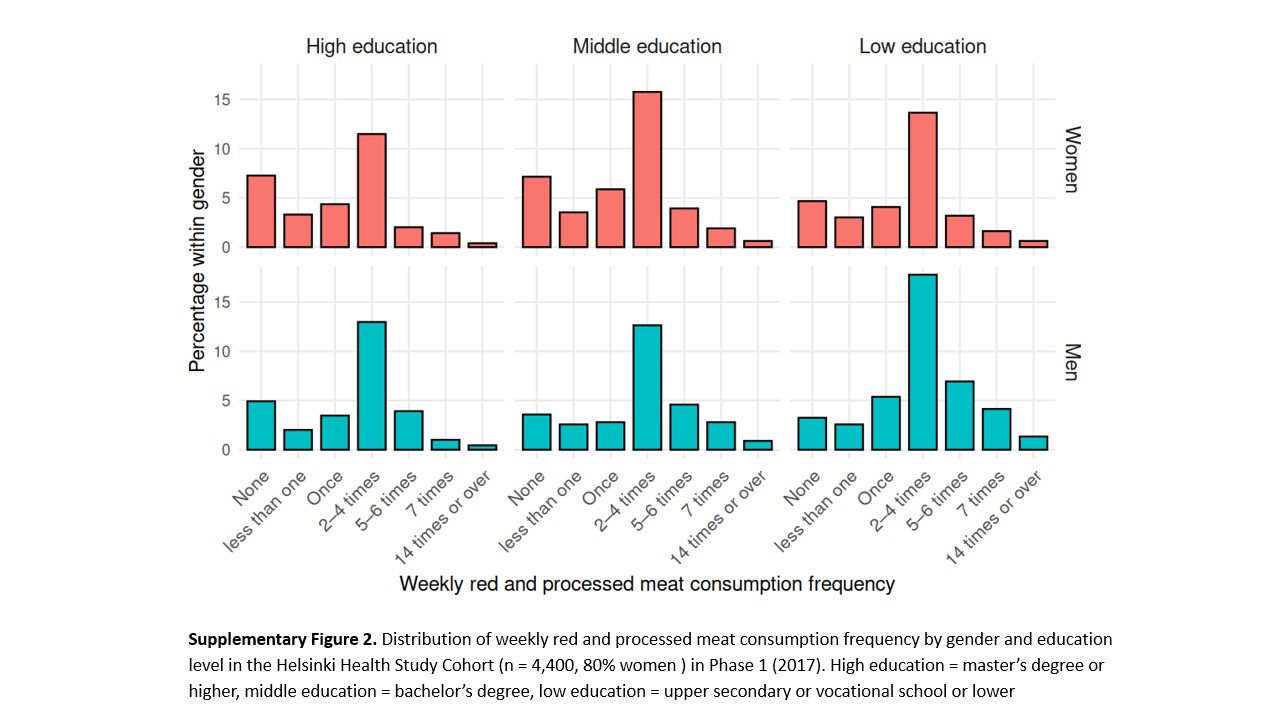
